# Prognostic value of sTREM-1 in COVID-19 patients: a biomarker for disease severity and mortality

**DOI:** 10.1101/2020.09.22.20199703

**Authors:** Pedro V. da Silva Neto, Jonatan C. S. de Carvalho, Vinícius E. Pimentel, Malena M. Pérez, Ingryd Carmona-Garcia, Nicola T. Neto, Diana M. Toro, Camilla N. S. Oliveira, Thais F. C. Fraga-Silva, Cristiane M. Milanezi, Lilian C. Rodrigues, Cassia F. S. L. Dias, Ana C. Xavier, Giovanna S. Porcel, Isabelle C. Guarneri, Kamila Zaparoli, Caroline T. Garbato, Jamille G. M. Argolo, Ângelo A. F. Júnior, Alessandro P. de Amorim, Augusto M. Degiovani, Dayane P. da Silva, Debora C. Nepomuceno, Rafael C. da Silva, Leticia F. Constant, Fátima M. Ostini, Marley R. Feitosa, Rogerio S. Parra, Fernando C. Vilar, Gilberto G. Gaspar, José J. R. da Rocha, Omar Feres, Rita C. C. Barbieri, Fabiani G. Frantz, Sandra R. Maruyama, Elisa M. S. Russo, Angelina L. Viana, Ana P. M. Fernandes, Isabel K. F. M. Santos, Vânia L. D. Bonato, Marcelo Dias-Baruffi, Adriana Malheiro, Ruxana T. Sadikot, Cristina R. B. Cardoso, Lúcia H. Faccioli, Carlos A. Sorgi

## Abstract

**Background:** The uncontrolled inflammatory response plays a critical role in the novel coronavirus disease (COVID-19) and triggering receptor expressed on myeloid cells-1 (TREM-1) is thought to be intricate to inflammatory signal amplification. This study aims to investigate the association between soluble TREM-1 (sTREM-1) and COVID-19 as a prognostic biomarker to predict the disease severity, lethality and clinical management.

**Methods:** We enrolled 91 patients with COVID-19 in domiciliary care (44 patients) or in hospital care (47 patients), who were classified after admission into mild, moderate, severe and critical groups according to their clinical scores. As non-COVID-19 control, 30 healthy volunteers were included. Data on demographic, comorbidities and baseline clinical characteristics were obtained from their medical and nurse records. Peripheral blood samples were collected at admission and after hospitalization outcome to assess cytokine profile and sTREM-1 level by specific immunoassays.

**Results:** Within COVID-19 patients, the highest severity was associated with the most significant elevated plasma levels sTREM-1. Using receiver operating curve analysis (ROC), sTREM-1 was found to be predictive of disease severity (AUC= 0.988) and the best cut-off value for predicting in-hospital severity was ≥ 116.5 pg/mL with the sensitivity for 93.3% and specificity for 95.8%. We also described the clinical characteristics of these patients and explored the correlation with markers of the disease aggravation. The levels of sTREM-1 were positively correlated with IL-6, IL-10, blood neutrophils counts, and critical disease scoring (r= 0.68, *p*<0.0001). On the other hand, sTREM-1 level was significantly negative correlated with lymphocytes counting, and mild disease (r= −0.42, *p*<0.0001). Higher levels of sTREM-1 were related to poor outcome and death, patients who received dexamethasone tended to have lower sTREM-1 levels.

**Conclusion:** Our results indicated that sTREM-1 in COVID-19 is associated with severe disease development and a prognostic marker for mortality. The use of severity biomarkers such as sTREM-1 together with patients clinical scores could improve the early recognition and monitoring of COVID-19 cases with higher risk of disease worsening.

## Introduction

The new coronavirus disease (COVID-19), caused by severe acute respiratory syndrome coronavirus 2 (SARS-CoV-2), it became a worldwide public health problem due to high rates of morbidity and mortality. Approximately 80% of patients infected with SARS-CoV-2 may have mild symptoms or are asymptomatic. However, the virus is capable of causing serious clinical manifestations, such as acute respiratory distress syndrome (ARDS), cardiovascular and coagulopathy disorders and shock; resulting in multi-organ system dysfunction in some patients (1–10). Patients with older ages, male gender, obesity, and chronic comorbidities such as cardiovascular disease, diabetes, chronic respiratory disease, and cancer were more likely to have worse outcomes (11–13). Moreover, patients with COVID-19 present multiple hematological abnormalities, of which lymphopenia and thrombocytopenia were prominent (14).

Similar to SARS, inflammation plays a major role in the pathophysiology of COVID-19 and patients exhibiting severe disease may have systemic high concentrations of inflammatory makers in serum samples, such as IL-6, TNF-α, C-reactive protein (CRP) and D-dimer (10,15–21). Pathogen infections generate an innate response that is initiated by the recognition of pathogen-associated molecular patterns (PAMPs) by pattern recognition receptors (PRRs) followed an antigen-specific adaptive immune response. Both innate and adaptive immune responses induce the activation of cell receptors and soluble mediators that trigger inflammation and its balance is critical for controlling tissue damage (22). Significant evidence indicates that a dysregulated immune and inflammatory response contributes to the clinical presentation of patients with severe COVID-19 disease.

Triggering receptor expressed on myeloid cells 1 (TREM-1) is a member of the immunoglobulin superfamily expressed in myeloid and epithelial cells. TREM-1 activation induces secretion of TNF-α, IL-6, IL-1β, IL-2, IL-12p40 by monocytes, macrophages and dendritic cells, enhancing inflammation during infections by different pathogens, namely Influenza A virus, Dengue virus, Hepatitis C virus, *Plasmodium falciparum, Staphylococcus aureus* and *Pseudomonas aeruginosa* (23–29). Immune responses against to virus and bacterial are modulated by activation of TREM-1 in macrophages. TREM-1 KO mice are protected from severe influenza although viral clearance is unaltered (30). Similarly, HIV infection, or even exposure to the HIV proteins, Tat or gp120, in the absence of infection (31) induce expression of TREM-1 suggesting that TREM-1 may have a pathogenic role in viral infections and immune response.

Soluble form of the TREM-1 (sTREM-1) has been reported in plasma samples during infection and inflammation (32–35). The sTREM-1 is a 27kDa polypeptide in wich the with the extracellular domain is released from the cell surface by action of metalloproteinases (36). Interestingly, the serum sTREM-1 level measurements could be a valuable diagnostic biomarker and may be useful in predicting severe prognosis in diseases related to inflammatory processes, as seen in septic patients, Hepatitis B virus, Human Immunodeficiency virus (HIV), Crimean Congo Hemorrhagic Fever and *Mycobacterium tuberculosis infections* (27,33,37–39).

Given that SARS-CoV-2 infection induces a hyper-inflammatory response, we hypothesized that TREM-1 activation in the COVID-19 illness triggers upregulation and shedding into sTREM-1. Because, sTREM-1 has been validated as prognostic markers in various inflammatory disorders, we performed herein a prospective study to investigate the prognostic utility of sTREM-1 in patients with COVID-19, as an important biomarker that can predicts disease severity and qualify effective interventions for managing patients that need intensive supportive care upon hospitalization.

## MATERIAL AND METHODS

### Study Design and subjects

This prospective study was conduced at *Hospital Santa Casa de Misericórdia of Ribeirão Preto* and *Hospital São Paulo of Ribeirão Preto - Brazil* from April to August of 2020 and was conducted with strict and reasonable inclusion and exclusion criteria. The patient inclusion criteria were adults (aged over 18 years), positive for SARS-CoV-2 infection that understood and agreed to enroll in this study. Controls (healthy adult volunteers) were tested negative for SARS-CoV-2. The exclusion criteria were individuals younger than 18 years of age and pregnant or lactating women. In total, 30 control subjects were enrolled at the beginning of the COVID-19 pandemic with a negative test for SARS-CoV-2 and 91 patients with a positive test for SARS-CoV-2, using genomic RNA assay performed in a nasopharyngeal/oropharyngeal swab RT-PCR (Kit Biomol OneStep/COVID-19) or serology specific IgM and IgG antibodies (SARS-CoV-2 antibody test^®^, Guangzhou Wondfo Biotech, China).

### Data collection

Members of our team carefully collected and reviewed the medical records of each patient. The data were searched and compiled from the electronic medical and nursering record systems. We included sociodemographic information, comorbidities, medical history, clinical symptoms, routine laboratory tests, immunological tests, chest computed tomography (CT) scans results, clinical interventions, and outcomes. These informations were documented on a standardized record form. Data collection of laboratory results was defined using the first-time examination at admission (within 24 h after admission), defined as primary endpoint, and the secondary endpoint was clinical outcome (death or recovery).

### Severity assessment

Clinically, severity of the COVID-19 patients was classified into mild, moderate, severe, and critical ill according to the statement in the Novel Coronavirus Pneumonia Diagnosis and Treatment Guideline (7th ed.), with modifications (40). The clinical manifestations were categorized as: i) mild group - positive for SARS-CoV-2 infection, with or without the following symptoms: diarrhea, cough, fever, headache, loss of taste (ageusia)/ smell (anosmia), myalgia, nausea, and vomiting, oxygen saturation between 94-99 % on room air; ii) moderate group - patients having manifestation of mild disease, with symptoms including dyspnea, oxygen saturation ≥ 93% on room air and arterial partial pressure of oxygen (PaO_2_)/ oxygen concentration (FiO_2_) between 250-300 mmHg, do not need invasive ventilation, allegedly using nasal-cannula oxygen (2-4 L/min) or oxygen masks ventilation (4-12 L/min); iii) severe group - possible admission to intensive-care units, severe respiratory distress, oxygen saturation < 93% on room air and PaO_2_/ FiO_2_ < 250mmHg, need non-invasive ventilation, such as oxygen masks ventilation (10-15 L/min); and iv) critical group - admission to intensive-care units, acute respiratory distress syndrome, need invasive ventilation, PaO_2_/ FiO_2_ <200mmHg, with or without one or more additional parameters: need hemodialysis, sepsis, septic shock, and multiorgan dysfunction (41).

### Laboratory Methods

Blood samples were collected by venipuncture in tubes with vacuum collection system. Two tubes (5mL capacity) were collected from each patient: one tube containing EDTA anticoagulant (BD Vacutainer^®^ EDTA K2 Franklin Lakes, New Jersey, USA) to perform hematological tests, and a tube containing heparin anticoagulant (BD SST^®^ Gel Advance^®^ Franklin Lakes, New Jersey, USA) to obtain plasma used to quantify levels of circulating protein mediators by flow cytometry or ELISA assay. Plasma was separated from whole blood samples and stored in a -80°C freezer. The clinical laboratory investigation included a complete blood count and biochemical serum tests (including liver and kidney function), as well as the determination of the coagulation factors and myocardial enzyme spectrum.

### Cytokines measurement

The cytokines IL-6 and IL-10 were quantified by the Cytometric Bead Array technique, according to the manufacturer’s specifications (BD™Human Inflammatory Cytokine CBA Kit, Catalog No. 551811, Lot: 9341655). Briefly, 50μl of plasma samples were mixed with capture antibody-coupled beads, and then 50μl fluorescent labeled (PE) detection antibodies. The samples were mixed and incubated at room temperature in the dark. After 3 h incubation, the beads were washed and resuspended with PBS 1% paraformaldehyde (*v/m –* Sigma Chemical Co. St. Louis, MO – USA). A recombinant protein standard for each cytokine that provides an internal control was included. The detection range of each cytokine was 2.5 pg/ml to 5000 pg/ml. Data acquisition was performed using a FACSCanto II flow cytometer (FACS CantoTMII – BD Biosciences, San Diego, CA, USA). Data was analyzed using FCAP Array 3.0 software and expressed as mean fluorescence intensity (MIF) for each serum cytokine (BD Biosciences, San Diego, CA, USA).

### Serum sTREM-1 assay

Circulating level of sTREM-1 were measured in plasma by an established available enzyme linked immunosorbent assay kit (DuoSet-Human TREM-1, R&D System, Minneapolis, USA), with a detection range of 93.8 – 6,000 pg/mL. The methodology was in according to the protocol described by the manufacturer (R&D Systems, Minneapolis, MN).

### Computation of Correlation Matrix

The dependence of multiple variables: sTREM-1 release (pg/mL); oxygen saturation (sO_2_ %); age (years); neutrophil count (x 10^9^ cells/L); lymphocytes count (x 10^9^ cells/L); hypertension (systole); glycemia (mg/dL); male gender; clinical scores (mild, moderate, severe and critical); IL-6 (pg/mL) and IL-10 amount (pg/mL); was calculated using Pearson correlation with cor() function in R (version 4.0.2) through RStudio (version 1.1.463). The computed correlation matrix was graphically displayed using corrplot function from R package “corrplot” (Version 0.84) (42). Correlation coefficients (r) and *p*-values of the correlation matrix were formatted and tabulated using Hmisc R package (43) and the values available at Supplemental Table S2.

### Statistical analysis

Data is presented as either counts or percentages (%) (for categorical) or median (IQR) and standard deviations (SD) for continuous data. Intergroup comparisons were made by the Mann-Whitney *U* test (non-normally distributed continuous variables) and chi-square test or Fischer’s exact test for categorical variables. One-way analysis of variance nonparametric test (Kruskal–Wallis test) was used for multiple comparisons. The accuracy of predictor was determined by area under the receiver operating characteristic (ROC) curve (AUC). The AUCs, with 95% confidence intervals, were computed to assess the diagnostic values of sTREM-1; AUCs > 0.70 were considered clinically relevant. Comparative analysis between groups was performed using GraphPad Prism™software (version 8.4.2) (San Diego, CA, USA). Statistical test differences were considered significant if the *p* values were < 0.05.

### Ethical approval

The procedures followed in this study were approved by the institutional ethics board of *Faculdade de Ciências Farmacêuticas de Ribeirão Preto* – *Universidade de São Paulo* and Brazil National Ethics Committee (CONEP, CAAE: 30525920.7.000.5403). Written informed consent was obtained from recruited participants of the study.

## RESULTS

### Demographic data, initial clinical signs and laboratory characteristics of study subjects

In total, 91 patients with laboratory-confirmed COVID-19 and 30 healthy volunteers were recruited to the study (Table 1). In this study population, 44 patients were diagnosed with mild disease with residential care and 47 patients with severe disease with hospital care on admission. The median age was 32.0 years (range, 23–80) for healthy volunteers and 49.0 years (range, 18–94) for all patients infected with SARS-CoV-2. The average age was significantly higher in the hospitalized patients than in residential care patients (Table 1). There was significant difference in the female ratio between the healthy control group and hospitalized COVID-19 patients (Table 1). Most patients had comorbidities or coexisting disorders, including hypertension (29.6%), heart disease (10.9%), diabetes (18.6%), neurological disorders (6.5%), cancer (5.4%), history of smoking (19.7%), and history of stroke (3.2%). The number of comorbidities was significantly higher in COVID-19 patients than in control group or in hospitalized patients compared to residential care subjects (Table 1).

**Table 1.**
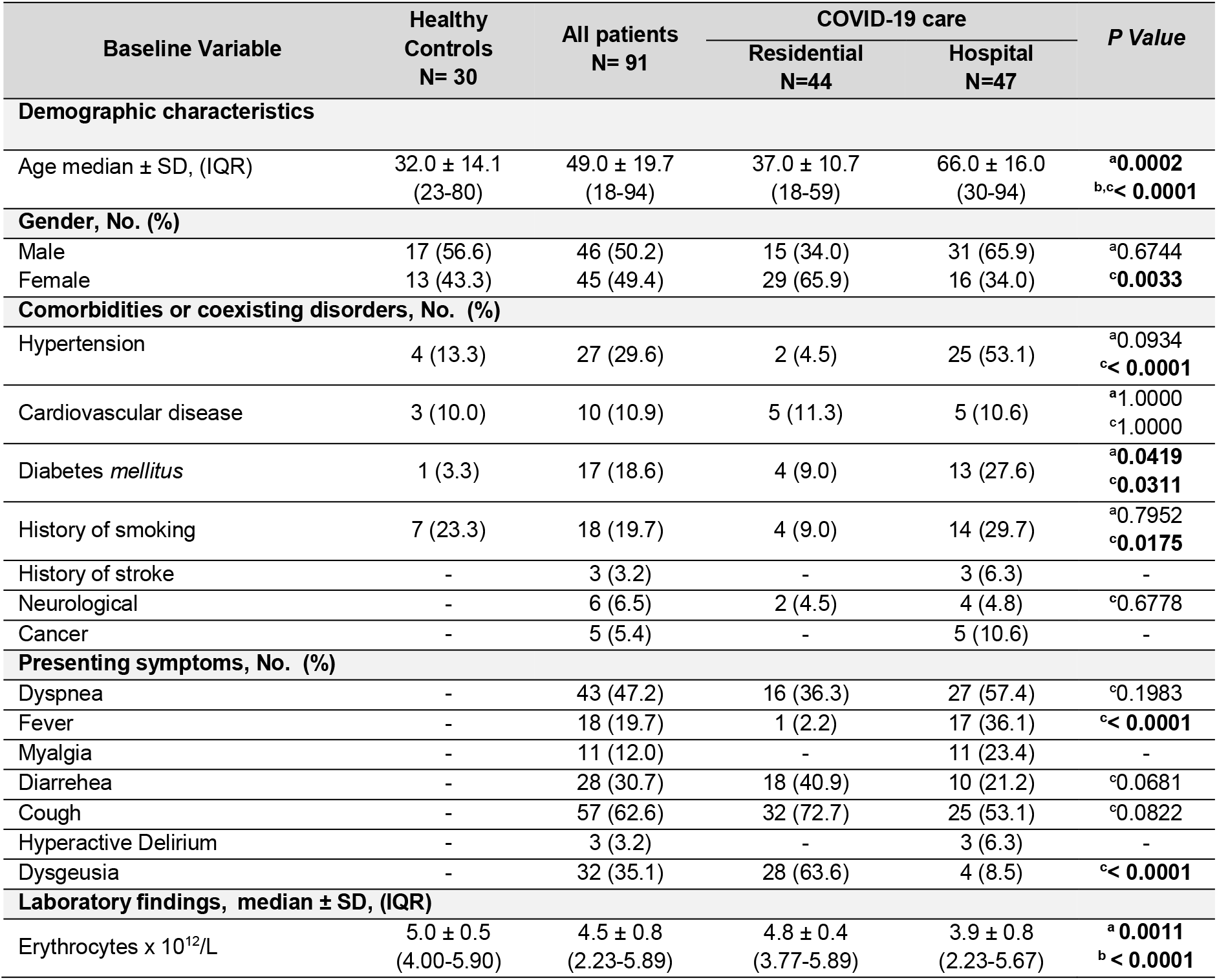
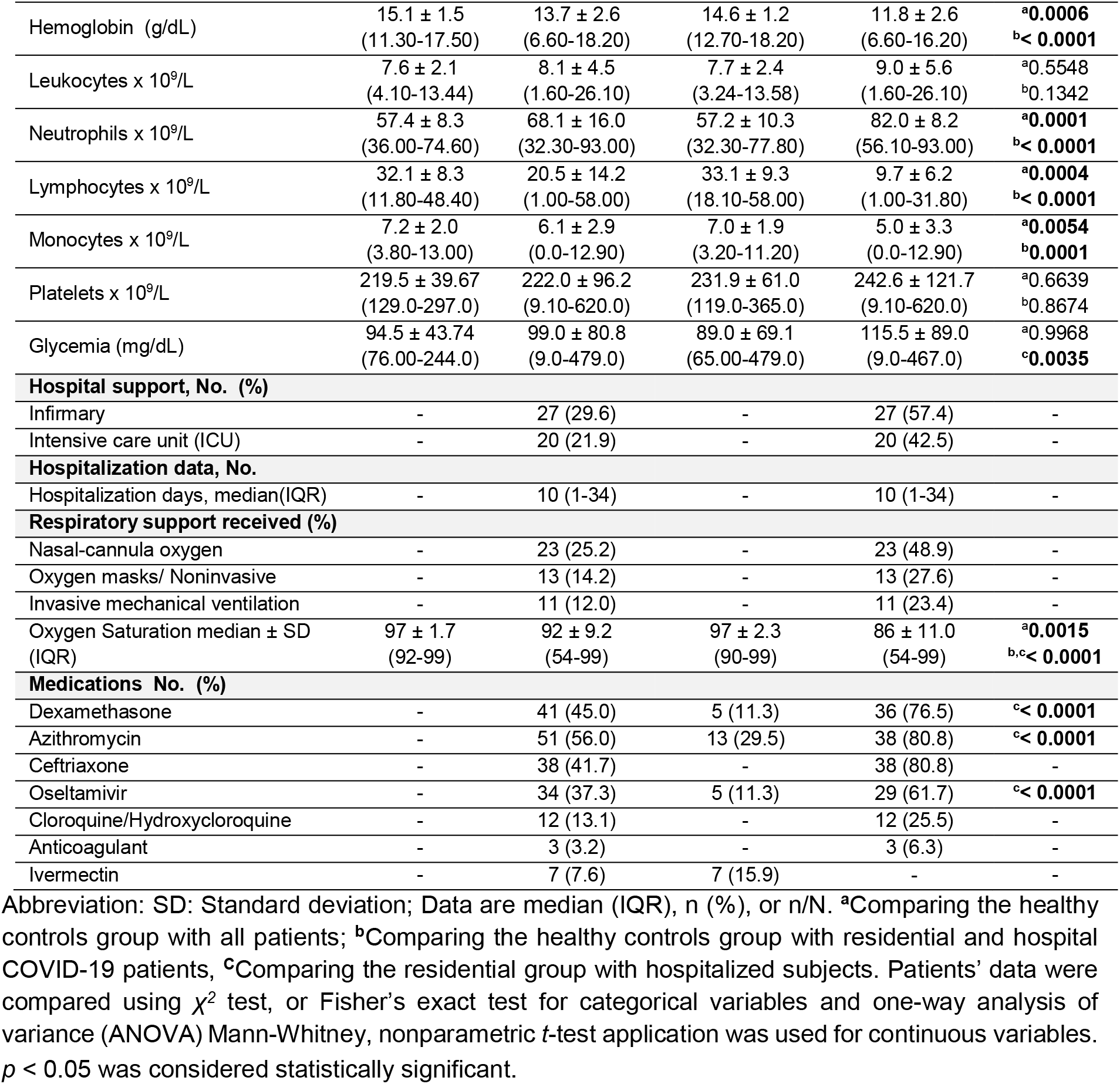
Demographics and clinical characteristics of patients infected with SARS-CoV-2.

| Baseline Variable | Healthy Controls<br>N= 30 | All patients<br>N= 91 | COVID-19 care |  | P Value |
| --- | --- | --- | --- | --- | --- |
|  |  |  | Residential<br>N=44 | Hospital<br>N=47 |  |
| Demographic characteristics |  |  |  |  |  |
| Age median ± SD, (IQR) | 32.0 ± 14.1<br>(23-80) | 49.0 ± 19.7<br>(18-94) | 37.0 ± 10.7<br>(18-59) | 66.0 ± 16.0<br>(30-94) | <sup>a</sup>**0.0002**<br><sup>b,c</sup>**< 0.0001** |
| Gender, No. (%) |  |  |  |  |  |
| Male | 17 (56.6) | 46 (50.2) | 15 (34.0) | 31 (65.9) | <sup>a</sup>0.6744 |
| Female | 13 (43.3) | 45 (49.4) | 29 (65.9) | 16 (34.0) | <sup>c</sup>**0.0033** |
| Comorbidities or coexisting disorders, No. (%) |  |  |  |  |  |
| Hypertension | 4 (13.3) | 27 (29.6) | 2 (4.5) | 25 (53.1) | <sup>a</sup>0.0934<br><sup>c</sup>**< 0.0001** |
| Cardiovascular disease | 3 (10.0) | 10 (10.9) | 5 (11.3) | 5 (10.6) | <sup>a</sup>1.0000<br><sup>c</sup>1.0000 |
| Diabetes *mellitus* | 1 (3.3) | 17 (18.6) | 4 (9.0) | 13 (27.6) | <sup>a</sup>**0.0419**<br><sup>c</sup>**0.0311** |
| History of smoking | 7 (23.3) | 18 (19.7) | 4 (9.0) | 14 (29.7) | <sup>a</sup>0.7952<br><sup>c</sup>**0.0175** |
| History of stroke | - | 3 (3.2) | - | 3 (6.3) | - |
| Neurological | - | 6 (6.5) | 2 (4.5) | 4 (4.8) | <sup>c</sup>0.6778 |
| Cancer | - | 5 (5.4) | - | 5 (10.6) | - |
| Presenting symptoms, No. (%) |  |  |  |  |  |
| Dyspnea | - | 43 (47.2) | 16 (36.3) | 27 (57.4) | <sup>c</sup>0.1983 |
| Fever | - | 18 (19.7) | 1 (2.2) | 17 (36.1) | <sup>c</sup>**< 0.0001** |
| Myalgia | - | 11 (12.0) | - | 11 (23.4) | - |
| Diarrhea | - | 28 (30.7) | 18 (40.9) | 10 (21.2) | <sup>c</sup>0.0681 |
| Cough | - | 57 (62.6) | 32 (72.7) | 25 (53.1) | <sup>c</sup>0.0822 |
| Hyperactive Delirium | - | 3 (3.2) | - | 3 (6.3) | - |
| Dysgeusia | - | 32 (35.1) | 28 (63.6) | 4 (8.5) | <sup>c</sup>**< 0.0001** |
| Laboratory findings, median ± SD, (IQR) |  |  |  |  |  |
| Erythrocytes x 10<sup>12</sup>/L | 5.0 ± 0.5<br>(4.00-5.90) | 4.5 ± 0.8<br>(2.23-5.89) | 4.8 ± 0.4<br>(3.77-5.89) | 3.9 ± 0.8<br>(2.23-5.67) | <sup>a</sup>**0.0011**<br><sup>b</sup>**< 0.0001** |
| Hemoglobin (g/dL) | 15.1 ± 1.5<br>(11.30-17.50) | 13.7 ± 2.6<br>(6.60-18.20) | 14.6 ± 1.2<br>(12.70-18.20) | 11.8 ± 2.6<br>(6.60-16.20) | <sup>a</sup>**0.0006**<br><sup>b</sup>**< 0.0001** |
| Leukocytes x 10<sup>9</sup>/L | 7.6 ± 2.1<br>(4.10-13.44) | 8.1 ± 4.5<br>(1.60-26.10) | 7.7 ± 2.4<br>(3.24-13.58) | 9.0 ± 5.6<br>(1.60-26.10) | <sup>a</sup>0.5548<br><sup>b</sup>0.1342 |
| Neutrophils x 10<sup>9</sup>/L | 57.4 ± 8.3<br>(36.00-74.60) | 68.1 ± 16.0<br>(32.30-93.00) | 57.2 ± 10.3<br>(32.30-77.80) | 82.0 ± 8.2<br>(56.10-93.00) | <sup>a</sup>**0.0001**<br><sup>b</sup>**< 0.0001** |
| Lymphocytes x 10<sup>9</sup>/L | 32.1 ± 8.3<br>(11.80-48.40) | 20.5 ± 14.2<br>(1.00-58.00) | 33.1 ± 9.3<br>(18.10-58.00) | 9.7 ± 6.2<br>(1.00-31.80) | <sup>a</sup>**0.0004**<br><sup>b</sup>**< 0.0001** |
| Monocytes x 10<sup>9</sup>/L | 7.2 ± 2.0<br>(3.80-13.00) | 6.1 ± 2.9<br>(0.0-12.90) | 7.0 ± 1.9<br>(3.20-11.20) | 5.0 ± 3.3<br>(0.0-12.90) | <sup>a</sup>**0.0054**<br><sup>b</sup>**0.0001** |
| Platelets x 10<sup>9</sup>/L | 219.5 ± 39.67<br>(129.0-297.0) | 222.0 ± 96.2<br>(9.10-620.0) | 231.9 ± 61.0<br>(119.0-365.0) | 242.6 ± 121.7<br>(9.10-620.0) | <sup>a</sup>0.6639<br><sup>b</sup>0.8674 |
| Glycemia (mg/dL) | 94.5 ± 43.74<br>(76.00-244.0) | 99.0 ± 80.8<br>(9.0-479.0) | 89.0 ± 69.1<br>(65.00-479.0) | 115.5 ± 89.0<br>(9.0-467.0) | <sup>a</sup>0.9968<br><sup>c</sup>**0.0035** |
| **Hospital support, No. (%)** |  |  |  |  |  |
| Infirmary | - | 27 (29.6) | - | 27 (57.4) | - |
| Intensive care unit (ICU) | - | 20 (21.9) | - | 20 (42.5) | - |
| **Hospitalization data, No.** |  |  |  |  |  |
| Hospitalization days, median(IQR) | - | 10 (1-34) | - | 10 (1-34) | - |
| **Respiratory support received (%)** |  |  |  |  |  |
| Nasal-cannula oxygen | - | 23 (25.2) | - | 23 (48.9) | - |
| Oxygen masks/ Noninvasive | - | 13 (14.2) | - | 13 (27.6) | - |
| Invasive mechanical ventilation | - | 11 (12.0) | - | 11 (23.4) | - |
| Oxygen Saturation median ± SD (IQR) | 97 ± 1.7<br>(92-99) | 92 ± 9.2<br>(54-99) | 97 ± 2.3<br>(90-99) | 86 ± 11.0<br>(54-99) | <sup>a</sup>**0.0015**<br><sup>b,c</sup>**< 0.0001** |
| **Medications No. (%)** |  |  |  |  |  |
| Dexamethasone | - | 41 (45.0) | 5 (11.3) | 36 (76.5) | <sup>c</sup>**< 0.0001** |
| Azithromycin | - | 51 (56.0) | 13 (29.5) | 38 (80.8) | <sup>c</sup>**< 0.0001** |
| Ceftriaxone | - | 38 (41.7) | - | 38 (80.8) | - |
| Oseltamivir | - | 34 (37.3) | 5 (11.3) | 29 (61.7) | <sup>c</sup>**< 0.0001** |
| Cloroquine/Hydroxychloroquine | - | 12 (13.1) | - | 12 (25.5) | - |
| Anticoagulant | - | 3 (3.2) | - | 3 (6.3) | - |
| Ivermectin | - | 7 (7.6) | 7 (15.9) | - | - |

Abbreviation: SD: Standard deviation; Data are median (IQR), n (%), or n/N. <sup>a</sup>Comparing the healthy controls group with all patients; <sup>b</sup>Comparing the healthy controls group with residential and hospital COVID-19 patients, <sup>c</sup>Comparing the residential group with hospitalized subjects. Patients' data were compared using $\chi^2$ test, or Fisher's exact test for categorical variables and one-way analysis of variance (ANOVA) Mann-Whitney, nonparametric *t*-test application was used for continuous variables. *p* < 0.05 was considered statistically significant.

Generally, the most common initial symptoms in COVID-19 patients were cough (62.6%), dyspnea (47.2%) and dysgeusia (35.1%), followed by diarrhea, fever, muscle soreness, and hyperactive delirium (Table 1). In addition, the absolute number of erythrocytes, hemoglobin, neutrophils, lymphocytes and monocytes in the COVID-19 patients group were significant different than that in the control group, while the glycemic level was significant difference between the residential care and hospital care groups (Figure 1). The median time of hospitalization for COVID-19 patients was 10 days and some patients needed intensive care (42.5%). Also, those hospitalized patients received respiratory support: nasal-cannula oxygen (48.9%), oxygen masks ventilation (27,6%) and invasive mechanical ventilation (23.4%). The oxygen saturation was significantly lower in the hospitalized patients than in residential care patients (Table 1). The most common pharmacological treatments for COVID-19 patients were azithromycin (56.0%), dexamethasone (45.0%), ceftriaxone (41.7%), oseltamivir (37.3%), followed by cloroquine/hydroxycloroquine, ivermectin and anticoagulant.

**Figure 1.**
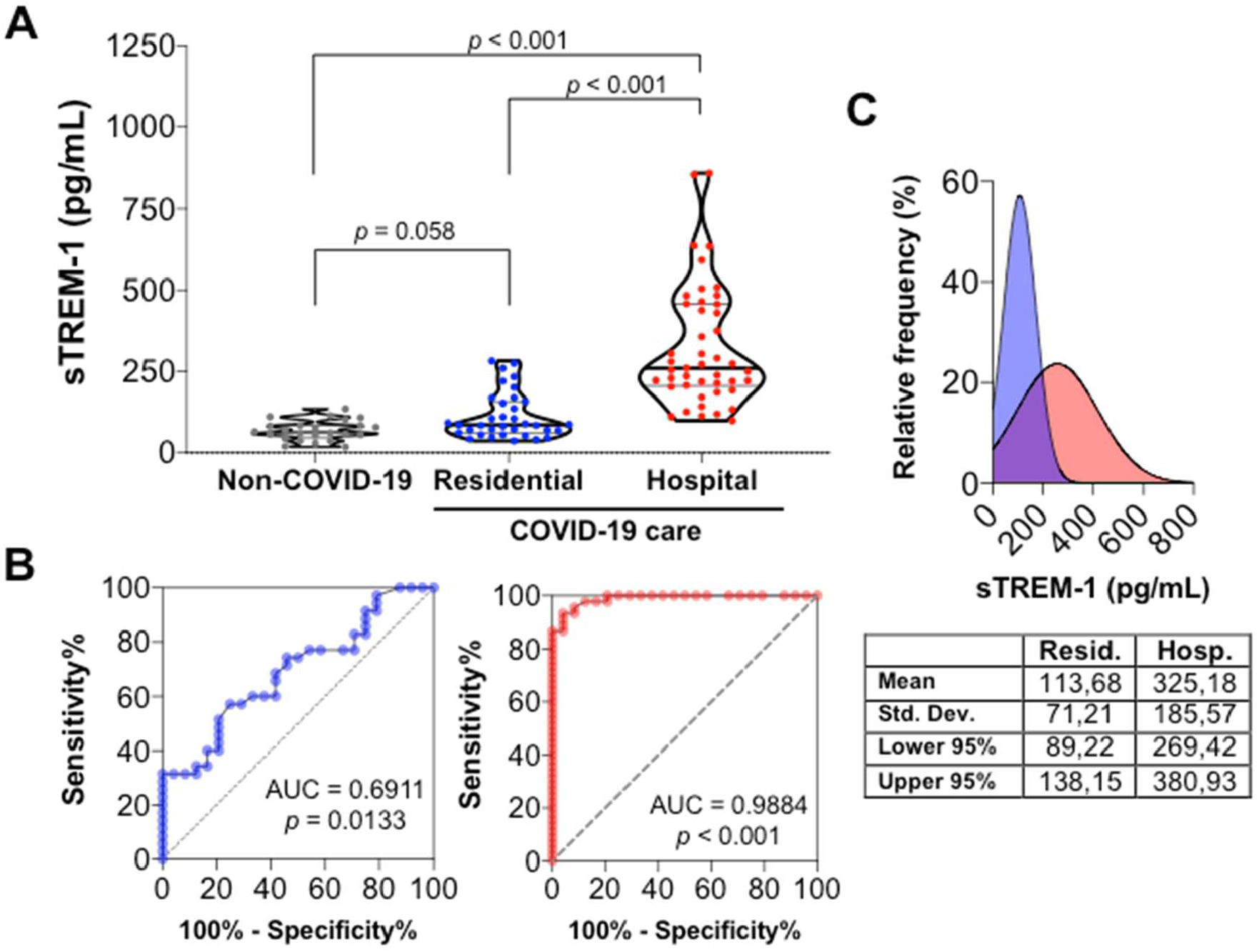
The values of sTREM-1 in COVID-19 patients are elevated. **(A)** The plasma concentration of sTREM-1 from COVID-19 patients with residential (n=44); or hospital care (n=47); and healthy controls (n=30) were analyzed and compared with each other. Median with range was presented. One-way analysis of variance nonparametric test (Kruskal–Wallis test) was used for multiple comparisons when data were not normally distributed. The differences between each group were presented with a *p* value in the graphic above the diagram. **(B)** ROC curves sTREM-1 for predicting disease on residential care (blue) and hospital care (red) patients with COVID-19. The area under the curve (AUC) and the comparative *p* value between COVID-19 patients and control group was designated in the graphic. **(C)** Relative frequency of residential care (blue) and hospital care (red) on sTREM-1 release in COVID-19 patients. The mean, standard deviation, CL lower 95% and CL upper 95% data of each group were presented in the table under the diagram.

### sTREM-1 release is significantly associated with disease severity

The potential role of sTREM-1 in regulating infectious diseases has been extensively investigated elsewhere. Here, we compared the levels of sTREM-1 in plasma samples from control group and COVID-19 patients, and also, assessed the correlation with severity illness. As shown in Figure 1A, sTREM-1 values were higher in patients with COVID-19 and significantly different in hospitalized group when compared with healthy volunteer controls, indicating an activation of immune response against SARS-CoV-2 infection. To evaluate the prognostic value of sTREM-1 in patients with COVID-19, ROC curves were drawn for patients with COVID-19 in residential and hospital care versus healthy controls.

The area under ROC curve of sTREM-1 for residential care patients, which represent mostly the mild disease, was proximal to the considered value for clinical relevance (AUC= 0.691); the preoperative concentration of ≥ 69.5 pg/mL was the optimal cutoff value for predicting COVID-19 (sensitivity = 71.43 (54.9– 83.6)%, specificity = 54.1%); the positive predictive values (PPV) of 91.6 (60.2– 98.7)% and the negative predictive values (NPV) of 48.9 (43.0–54.9)% were described (Figure 1B). However, the area under ROC curve of sTREM-1 for hospital care patients, which denote the severe disease, was higher for clinical relevance (AUC= 0.988); the sTREM-1 cut-off value of ≥ 116.5 pg/mL yielded sensitivity of 93.3 (82.1–97.7)%, specificity of 95.83%, the PPV of 97.5 (85.4– 99.6)%, and the NPV of 82.1 (66.7–91.4)% for differentiating patients with COVID-19 from those healthy subjects (Figure 1B). The plotted relative frequency of patients to sTREM-1 production, showed a high number of events for low level of sTREM-1 in residential care patients (mean = 113.68 pg/mL), and a spreader number of events for high levels of sTREM-1 in hospital care patients (mean = 325.18 pg/mL), considering a intersection phase between residential and hospitalized patients with COVID-19 (Figure 1C). Those results indicated the sTREM-1 as a potential marker predictive of COVID-19 severity illness.

### Correlation of sTREM -1 with disease severity, comorbidities and inflammatory mediators in COVID-19 patients

Subgroup analysis correlation of sTREM-1 in adverse outcome prediction was performed for patients according to different disease severity on admission. In this case, oxygen saturation (sO2), age, neutrophil count, lymphocytes count, hypertension, glycemia, male gender, clinical scores, IL-6 and IL-10 were identified as independent risk factors of adverse outcome in patients with COVID-19. For this, we performed analyses using the level of sTREM-1 in 91 patients’ plasma obtained during the period of SARS-CoV-2 infection diagnostic confirmation. In figure 2A, a correlation plot using correlation matrix (*r* values) showed statistically positively association of sTREM-1 level with critical disease, neutrophils count, age, and IL-10, IL-6 production. Conversely, we also observed a statistically negative association of sTREM-1 with mild disease and lymphocytes count. Full *r* and *p-*values of the correlation matrix are available in Supplemental Table S2. To further examine if higher sTREM-1 contributes to disease severity, we next analyzed sTREM-1 level in COVID-19 patients subgroups based on disease severity (Figure 2B). Despite significant differences between critical and severe or moderate and to mild groups, such significance was not seen for comparison between moderate and mild patients.

**Figure 2.**
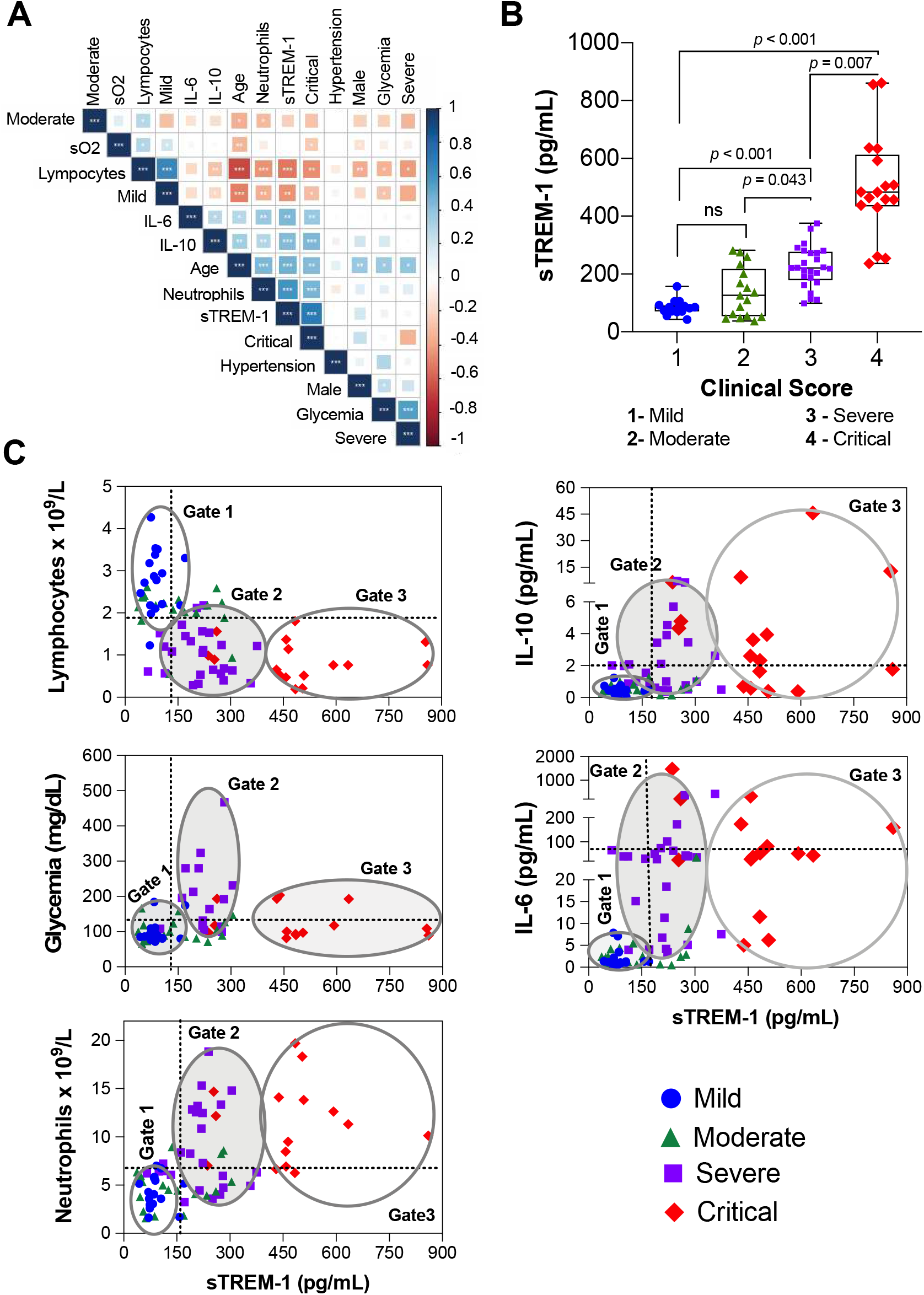
Clinical and immunological parameters strictly correlated with sTREM-1 levels in COVID-19 patients. **(A)** Correlogram of sTREM-1 and cytokine levels in the plasma of COVID-19 patients with comorbidities and clinical characteristics. Color scale sidebar indicates the correlation coefficients (r), where blue means positive correlation and red means negative correlation. Square size and color intensity are proportional to the correlation coefficients. Statistically significant correlations of pairwise variable comparisons are indicated by \**p*<0.05, \*\**p*<0.01 and \*\*\**p*<0.001. **(B)** The level of sTREM-1 in COVID-19 patients with different severity: mild (n= 23), moderate (n= 21), severe (n= 28) and critical (n= 19) group. Median with ranges was presented. One-way analysis of variance nonparametric test (Kruskal–Wallis test) was used for multiple comparisons when data were not normally distributed. The differences between each group were presented with a *p* value in the graphic above the diagram. **(C)** The correlation of plasma sTREM-1 and lymphocytes count, glycemia, neutrophils count, IL-10 and IL-6 amount in patients with COVID-19 was represented by a dot-plot graphic. The lines indicated the median value of different variables of all patients. Gates are placed around populations with common characteristics of patients with COVID-19.

As expected, in figure 2C we demonstrated a positive correlation between sTREM-1 and neutrophils count, IL-10 and IL-6 levels. However, when comparing these independent risk factors with the adverse outcome in patients with COVID-19, we observed a homogeneity values for mild and moderate disease (Gate 1), and variable increased levels for severe (Gate 2) and critical disease (Gate 3). On this way, we suggested that sTREM-1 value is a better predictor of clinical outcome of the disease than other risk factors or inflammatory mediators. In addition, sTREM-1 negatively correlated with lymphocyte count; and as assumed, sTREM-1 levels improved the discrimination between the mild/moderate to severe and critical group when compared to total lymphocytes number. Interesting, we observed that the glycemia index was higher in severe group than in mild/moderate and critical group, indicating that a single analysis of glycemia cannot be used to classify adequatelly all the COVID-19 severity ranges.

### Added Prognostic Value of sTREM -1 to predict mortality risk in COVID-19

To perform longitudinal analysis, sTREM-1 in patients with COVID-19 was further monitored during hospitalization. To better view the overall changes of sTREM-1 level, the plasma concentrations in the admission (baseline level) and after in the outcome treatment (death or recovery) were compared (Figure 3A). Of note, mostly patients treated with dexamethasone showed a reduced or stabilized sTREM-1 release after treatment, with only two-outcome recovery patient. In contrast, plasma sTREM-1 was significantly elevated in patients with COVID-19 who were not treated with dexamethasone. Notably, no recovery outcome was observed in these subjects and thus this analysis showed that sTREM-1 was a significant predictor of mortality.

**Figure 3.**
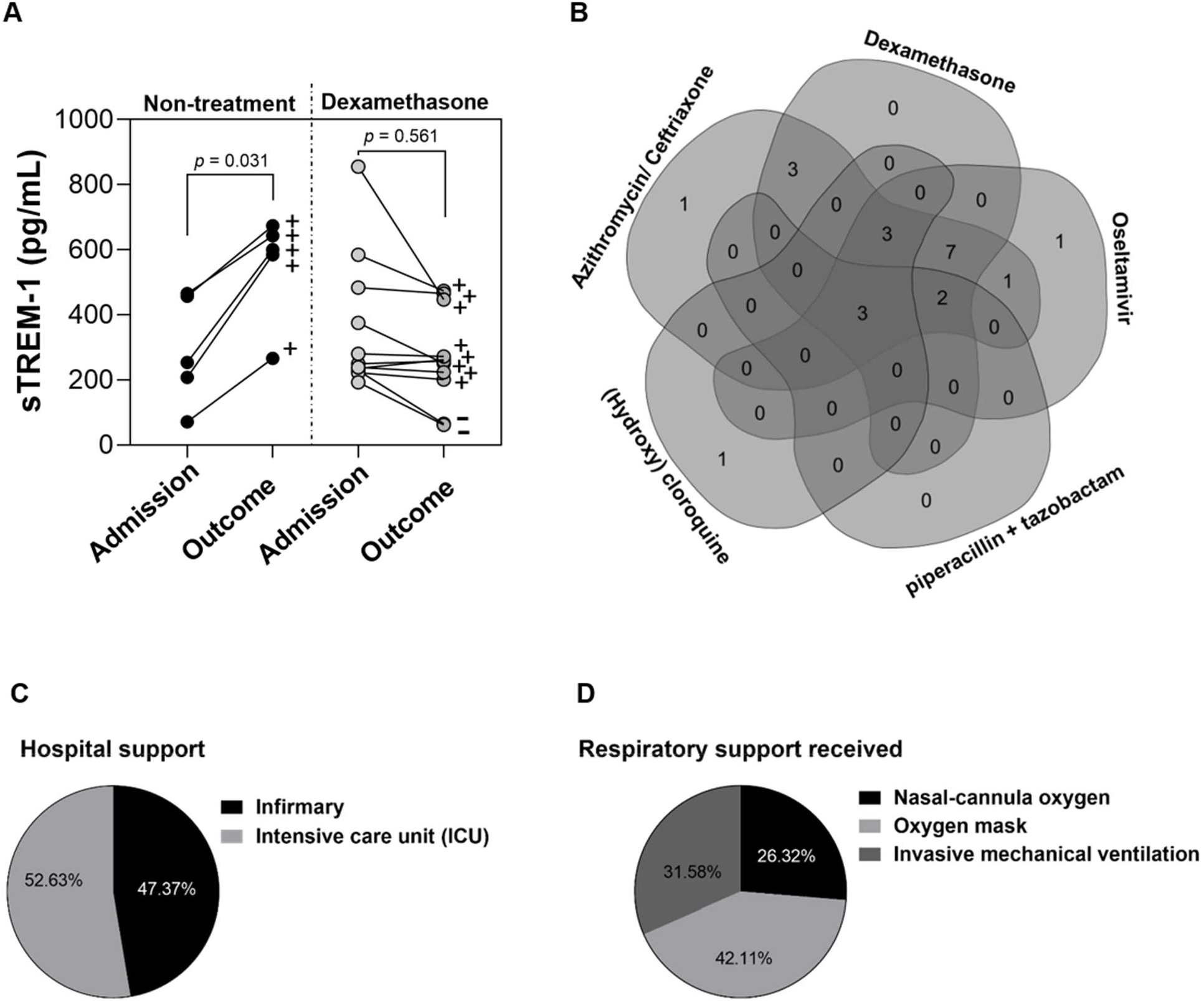
The kinetics of sTREM-1 in COVID-19 patients during hospitalization. **(A)** The plasma sTREM-1 level of critical patients during hospitalization was presented. The graphics indicated the individually value of sTREM-1 in the day of hospital-admission and after in the disease outcome, that can be death (+) or recovery (-). The individuals were segmented according with pharmacology treatment in dexamethasone-treatment and non-treatment with dexamethasone. Each circle indicates an individual patient. Nonparametric *t* test (Mann-Whitney U test) was used to data not normally distributed. The differences between the groups were presented with a *p* value in the graphics. **(B)** Venn diagram showed all pharmacological treatment relations between dexamethasone, Oseltamivir, Piperacillin+Tazobactam, Hydroxycloroquine/cloroquine, Azitromycin/Ceftriaxone of different sets patients with COVID-19 in hospital care. **(C)** Demonstrative percentages of patients in distinctive hospital and respiratory support during hospitalization.

In figure 3B, we showed our cohort of COVID-19 hospital pharmacological therapies. Although there are no approved treatments for COVID-19, the hospital protocols issued for antibiotics, antivirals and anti-inflammatory drugs. While treatment in this study was not stratified by severity of disease, 14 hospitalized patients in this cohort longitudinal analysis have been treated with dexamethasone (73.68%); 14 patients with antibiotics (azithromycin/ceftriaxone 73.68%), 12 patients with oseltamivir (63.15%), 5 patients with hydroxicloroquine/cloroquine (26.3%) and 1 patient treated with anticoagulant (5,2%). The combined pharmacology approach for these patients with COVID-19 was represented by a Venn diagram (Figure 3B). Changes in oxygen support level from hospital admission to recovery or death were showed in Figure 3C. In our cohort, 9 patients were in hospital infirmary care (47.3%) and 10 patients were in intensive care unit (52.63%). Among those patients, 6 were on invasive mechanical ventilation (31.5%), 8 on oxygen masks non-invasive ventilation (42.1%) and 5 on oxygen nasal-cannula (26.3%) during hospitalization. The details of hospital support and supportive therapies for patients with COVID-19 were demonstrated in Supplemental Table 1.

## DISCUSSION

To date, the most significant predictors of COVID-19 disease severity are relate to either activation or suppression of the host immune response (44,45). In the present study, we show for the first time in a population of patients in the city of Ribeirão Preto - Brazil with COVID-19 that plasma sTREM-1 concentrations might be a predictive factor and a biomarker for disease severity. Plasma sTREM-1 concentrations were gradually increased from mild to moderate, severe and critical forms of COVID-19. Indeed, ROC analysis confirms that sTREM-1 was an important predictor of disease progression in COVID-19 patients.

TREM-1 exhibit the ability to amplify proinflammatory innate immune response, such as by the Toll like receptors (TLR), which recognize a wide range of bacterial, fungal and viral components (46). In addition to neutrophils, monocytes and macrophages, TREM-1 is also expressed in epithelial and endothelial cells (25). Elevated sTREM-1 level are indicative of acute and chronic conditions including sepsis and pneumonia (47,48). Enhanced inflammatory response in macrophages was indicated as a mechanism by which TREM-1 signaling contributes to lung injury, suggesting that TREM-1 inhibition was a potential therapeutic target for neutrophilic lung inflammation and acute respiratory distress syndrome (ARDS) (49). Moreover, sTREM-1 level in sepsis could reflect an essential immune dysfunction, where excessive cleavage of TREM-1 could contribute to immunosuppression and death during a severe infection (50). Since severe infections can occur by multiple microorganism the broad prognostic of sTREM-1 implied its potential as a severity marker for all-cause febrile illness (51). However, fever was not the mostly common signs in admission patient with COVID-19 in our cohort, especially in mild and moderate disease. Thus, sTREM-1 has been recognized to be a valuable diagnostic and prognostic marker since it is easily detected using immunochemical assays. Nevertheless, the role of sTREM-1 in patients with COVID-19 remains unclear.

The number of COVID-19 patients is increasing dramatically worldwide and the early recognition of severe forms of COVID-19 is essential for timely triaging of patients. Some model analysis showed that IL-6 and CRP could be used as independent factors to predict the severity of COVID-19 (52). IL-6 is a multifunctional cytokine that presented a strong proinflammatory effect with functions in inflammation, tumor, and hematological diseases (53,54). Other work suggested that the predictive values of IL-10 and IL-6 should be preferentially evaluated for early diagnosis of patients with severe disease (18). IL-10 is highly abundant, especially during the adaptive immune response (55). Although, some studies pointed that plasma IL-6 and IL-10 levels could be used as a factor to predict the progression of COVID-19, in our cohort these values alone did not distinguish the severe and critical groups, but these cytokines were in lower levels in mild/moderate disease and higher expression was observed in some patients with severe/critical COVID-19. Patients with severe COVID-19 disease presented with increased leukocytosis, neutrophilia, lymphopenia, and thrombocytopenia than those with non-severe disease (56); and we also demonstrated a significant correlation between plasma sTREM-1 and those hematological parameters in severe COVID-19.

Instead, sTREM-1 levels were found to be independent discriminators of disease severity on admission, we also comprised that sTREM-1 could be an outcome predictor for patients with COVID-19. Therefore, markedly elevated plasma sTREM-1 levels in patients with COVID-19 might be the indication of excessive inflammatory response and contribute to severe/critical illness or even death. Generally, the most common therapeutic options for viral infections are directed at either blocking viral replication or modulation of immune response. Dexamethasone exhibits broad-spectrum anti-inflammatory effects. In patients hospitalized with COVID-19, the use of dexamethasone resulted in lower mortality (57). We observed a longitudinal sTREM-1detection in patient admission and outcome in hospital care; in patients with non-treatment with dexamethasone, sTREM-1 increased after admission and correlated with outcome death. However, patients that were treated with dexamethasone, in mostly the case, control and reduced the sTREM-1 production, but this phenomenon was not correlated with better recovery. It means a non-return point to anti-inflammatory treatment in COVID-19, and sTREM-1 value could expose that inflammatory status. It is likely that the beneficial effect of glucocorticoids in severe COVID-19 is subject on a selection of dose and time.

There are some limitations in our study. First, chronic diseases and secondary infection in some cases might exert effects on the increased plasma sTREM-1 level besides SARS-CoV-2 infection itself. Second, this study was limited by sample size. Larger studies with continuous monitoring of sTREM-1 are necessary to further confirm the prognostic effect in patients with severe COVID-19. Exacerbation of the situation is usually a dynamic process; thus the data at a certain time point may not accurately reflect change in the patient condition. Third, some patients with critical illness were not admitted to intensive care unit, which definitely had a negative impact on the outcome of those patients. Although our findings should be evaluated with larger number of samples by multi-center research, sTREM-1 is a potential marker of diagnostic to distinguish critical COVID-19 and poor-outcome, and monitoring the sTREM-1 levels will improve treatment efficacy and reduce mortality.

## CONCLUSION

In conclusion, our results suggested that admission plasma sTREM-1 levels play an important role in discriminating disease severity and predicting adverse outcome in patients with COVID-19. Patients with markedly elevated admission sTREM-1 should be provided more attention and supported treatment. In this perspective, we suggested the sTREM-1 value contribution to the clinical course of COVID-19 infection and highlight potential strategies for therapeutic intervention. Also, emerged as a possible prognostic biomarker readily detected in plasma of COVID-19 patients.

## Data Availability

Not applied

## ACKNOWLEDGEMENTS

The authors acknowledge the support of the ICU team of doctors, nurses, physiotherapists and the collaboration of *Hospital Santa Casa de Misericórdia of Ribeirão Preto* and *Hospital São Paulo of Ribeirão Preto*. The laboratory support from Supera Parque - Innovation and Technology Park-Ribeirão Preto/SP for testing infection by SARS-CoV-2 in healthy volunteers. The valuable contribution by Municipal Health Department of Ribeirão Preto city and Analysis Service Clinics (SAC) from Faculdade de Ciências Farmacêuticas de Ribeirão Preto – USP. We are grateful to Fabiana R. de Moraes for helping with Flow Cytometer analysis; and Professors Victor Hugo Aquino Quintana and Márcia Regina von Zeska Kress for sharing viral BSL2 lab.

## Supplementary Data

**Supplemental Table 1.**
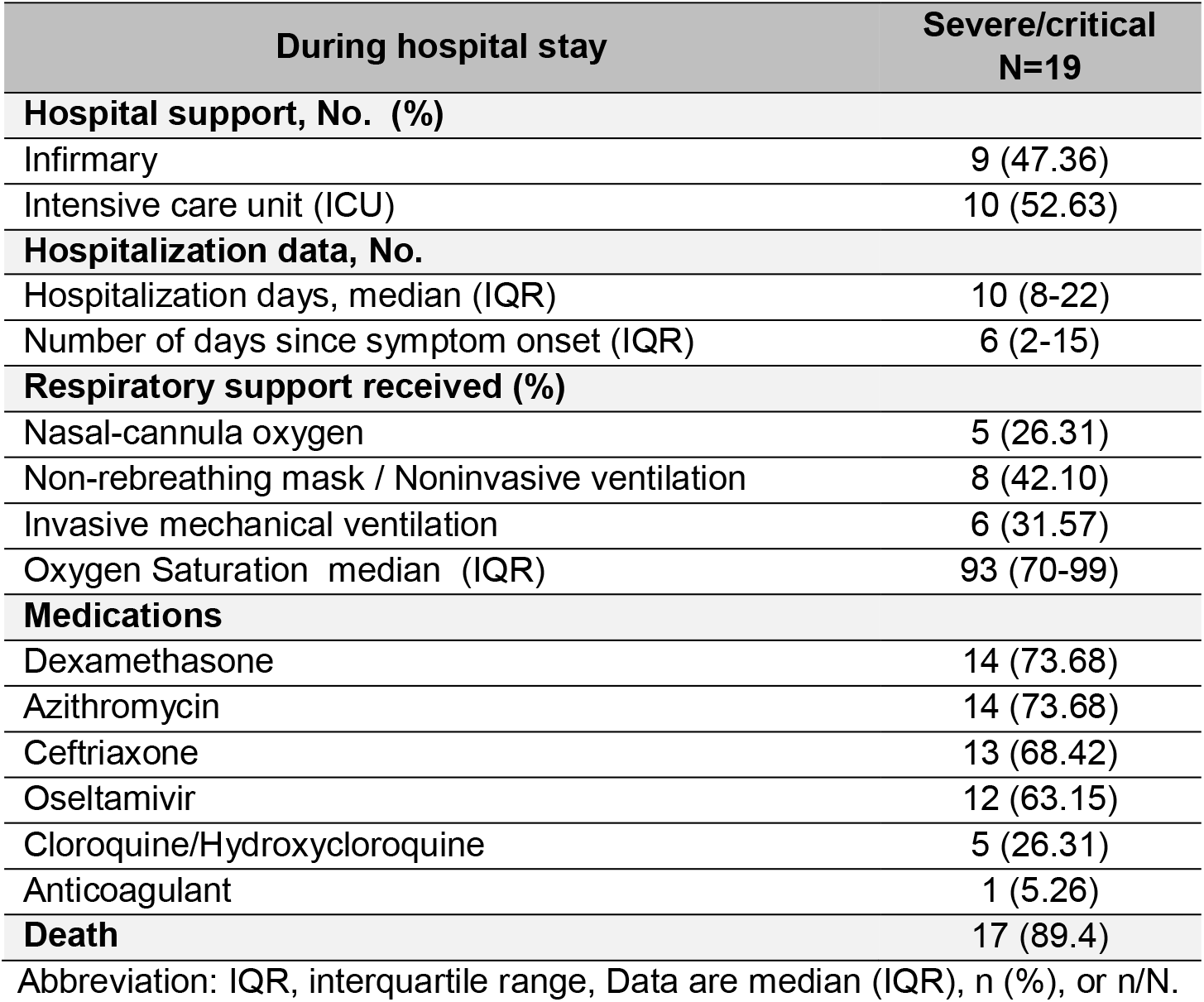
Hospital support, supportive therapies and medications of patients severe and critical infected with SARS-CoV-2.

**Supplemental Table 2.** Information about r and *p-*values of the correlation matrix of the graph shown in figure 2.

| Comparison | Row | Column | Correlation | p-value |
| --- | --- | --- | --- | --- |
| 1 | sTREM1 | Lymphocytes | -0,52170 | 1,14E-06 |
| 2 | sTREM1 | Neutrophils | 0,58800 | 1,88E-08 |
| 3 | Lymphocytes | Neutrophils | -0,44264 | 5,55E-05 |
| 4 | sTREM1 | Glycemia | 0,09286 | 0,42182 |
| 5 | Lymphocytes | Glycemia | -0,36861 | 0,00097 |
| 6 | Neutrophils | Glycemia | 0,09778 | 0,39753 |
| 7 | sTREM1 | IL10 | 0,42499 | 0,00012 |
| 8 | Lymphocytes | IL10 | -0,26048 | 0,02214 |
| 9 | Neutrophils | IL10 | 0,27028 | 0,01744 |
| 10 | Glycemia | IL10 | 0,12544 | 0,27703 |
| 11 | sTREM1 | IL6 | 0,43267 | 8,50E-05 |
| 12 | Lymphocytes | IL6 | -0,16860 | 0,14271 |
| 13 | Neutrophils | IL6 | 0,35654 | 0,00146 |
| 14 | Glycemia | IL6 | -0,04650 | 0,68803 |
| 15 | IL10 | IL6 | 0,27826 | 0,01427 |
| 16 | sTREM1 | Mild | -0,42227 | 0,00011 |
| 17 | Lymphocytes | Mild | 0,64575 | 2,28E-10 |
| 18 | Neutrophils | Mild | -0,34410 | 0,00218 |
| 19 | Glycemia | Mild | -0,26277 | 0,02095 |
| 20 | IL10 | Mild | -0,17832 | 0,12075 |
| 21 | IL6 | Mild | -0,14227 | 0,21709 |
| 22 | sTREM1 | Moderate | -0,26402 | 0,01871 |
| 23 | Lymphocytes | Moderate | 0,25992 | 0,02244 |
| 24 | Neutrophils | Moderate | -0,33536 | 0,00287 |
| 25 | Glycemia | Moderate | -0,23557 | 0,03917 |
| 26 | IL10 | Moderate | -0,17433 | 0,12942 |
| 27 | IL6 | Moderate | -0,14347 | 0,21321 |
| 28 | Mild | Moderate | -0,28445 | 0,01107 |
| 29 | sTREM1 | Severe | -0,04560 | 0,69176 |
| 30 | Lymphocytes | Severe | -0,40727 | 0,00026 |
| 31 | Neutrophils | Severe | 0,17927 | 0,12125 |
| 32 | Glycemia | Severe | 0,52996 | 8,52E-07 |
| 33 | IL10 | Severe | -0,02137 | 0,85461 |
| 34 | IL6 | Severe | -0,03733 | 0,74885 |
| 35 | Mild | Severe | -0,35237 | 0,00156 |
| 36 | Moderate | Severe | -0,36560 | 0,00100 |
| 37 | sTREM1 | Critical | 0,68800 | 6,59E-12 |
| 38 | Lymphocytes | Critical | -0,44213 | 8,02E-05 |
| 39 | Neutrophils | Critical | 0,45144 | 5,41E-05 |
| 40 | Glycemia | Critical | -0,05256 | 0,65648 |
| 41 | IL10 | Critical | 0,45644 | 4,36E-05 |
| 42 | IL6 | Critical | 0,37704 | 0,00093 |
| 43 | Mild | Critical | -0,27454 | 0,01639 |
| 44 | Moderate | Critical | -0,28493 | 0,01261 |
| 45 | Severe | Critical | -0,33736 | 0,00288 |
| 46 | sTREM1 | Male | 0,19819 | 0,08197 |
| 47 | Lymphocytes | Male | -0,39743 | 0,00038 |
| 48 | Neutrophils | Male | 0,15473 | 0,18202 |
| 49 | Glycemia | Male | 0,28389 | 0,01295 |
| 50 | IL10 | Male | 0,15893 | 0,17027 |
| 51 | IL6 | Male | -0,02913 | 0,80277 |
| 52 | Mild | Male | -0,21052 | 0,06431 |
| 53 | Moderate | Male | -0,17884 | 0,11721 |
| 54 | Severe | Male | 0,17882 | 0,11725 |
| 55 | Critical | Male | 0,20958 | 0,06921 |
| 56 | sTREM1 | Age | 0,43058 | 8,32E-05 |
| 57 | Lymphocytes | Age | -0,64649 | 2,83E-10 |
| 58 | Neutrophils | Age | 0,43821 | 7,52E-05 |
| 59 | Glycemia | Age | 0,33385 | 0,00321 |
| 60 | IL10 | Age | 0,33105 | 0,00349 |
| 61 | IL6 | Age | 0,27897 | 0,01467 |
| 62 | Mild | Age | -0,50132 | 2,94E-06 |
| 63 | Moderate | Age | -0,32776 | 0,00340 |
| 64 | Severe | Age | 0,41118 | 0,00020 |
| 65 | Critical | Age | 0,39176 | 0,00051 |
| 66 | Male | Age | 0,33291 | 0,00309 |
| 67 | sTREM1 | Systole | 0,12566 | 0,27942 |
| 68 | Lymphocytes | Systole | -0,10523 | 0,37222 |
| 69 | Neutrophils | Systole | -0,03093 | 0,79362 |
| 70 | Glycemia | Systole | 0,12893 | 0,27360 |
| 71 | IL10 | Systole | -0,11298 | 0,33785 |
| 72 | IL6 | Systole | -0,05190 | 0,66054 |
| 73 | Mild | Systole | -0,03689 | 0,75175 |
| 74 | Moderate | Systole | -0,05579 | 0,63217 |
| 75 | Severe | Systole | -0,04354 | 0,70879 |
| 76 | Critical | Systole | 0,11156 | 0,34401 |
| 77 | Male | Systole | -0,02091 | 0,85768 |
| 78 | Age | Systole | 0,02243 | 0,84853 |
| 79 | sTREM1 | SaO<sub>2</sub> | 0,02523 | 0,82987 |
| 80 | Lymphocytes | SaO<sub>2</sub> | 0,25259 | 0,02991 |
| 81 | Neutrophils | SaO<sub>2</sub> | -0,07563 | 0,52189 |
| 82 | Glycemia | SaO<sub>2</sub> | -0,03030 | 0,79774 |
| 83 | IL10 | SaO<sub>2</sub> | -0,02714 | 0,81847 |
| 84 | IL6 | SaO<sub>2</sub> | 0,03383 | 0,77480 |
| 85 | Mild | SaO<sub>2</sub> | 0,17241 | 0,13911 |
| 86 | Moderate | SaO<sub>2</sub> | 0,12471 | 0,28640 |
| 87 | Severe | SaO<sub>2</sub> | -0,07139 | 0,54555 |
| 88 | Critical | SaO<sub>2</sub> | -0,24686 | 0,03658 |
| 89 | Male | SaO<sub>2</sub> | -0,11772 | 0,31784 |
| 90 | Age | SaO<sub>2</sub> | -0,31813 | 0,00574 |
| 91 | Systole | SaO<sub>2</sub> | 0,03868 | 0,74523 |

## NOTES

### Author Contributions

PVSN, JCSC, CAS and conceptualization the study. PVSN, JCSC, VEP, NTN performed the main experimental procedure. MMP, supervised the collection of samples from patients. PVSN, JCSC, YCG, ACX, GSP, ICG, KZ, CT G, JGMA, AAFJ, help with samples collection. PVSN, JCSC, TFCFS, CMM, LCR, CFSLD, sample processed and fractionation. CNSO, YCG, TFCFS, CRBC, performed the main experimental cytokines measurement. SRM, analyzed the data by R program. PVSN, DMT, CAS, performed the statistical analysis of the data and wrote the article and prepared the figures. DPS, DCN, RCS, LFC, RCCB, APA, AMD, FMO, MRF, RSP, FCV, GGG, JJRR, OF, contributed to the collection of clinical specimens, demographic, helped clinical data management and clinical characteristics analysis from COVID-19 patients. PVSN, DMT, CAS, drafted the manuscript. All authors helped editing the manuscript. CAS and LHF supervised the project. FGF, VLDB, CRBC and LHF, provided study materials, reagents and other analysis tools and equipment. CAS, FGF, SRCM, EMSR, ALV, APMF, IKFMS, VLDB, CRBC, MDB, AM, RTS, LHF, coordination and idealization of the project; and critical revision of the manuscript for important intellectual concept. All authors have read and agreed to the published version of the manuscript.

